# In vivo measurement and prediction of cell-type-resolved non-coding variant effects in human immunity

**DOI:** 10.64898/2026.05.05.26352448

**Authors:** Qiaoqiao Liu, Jing Zhang, Yeping Zhu, Kangchun Wang, Changming Zhang, Jiahui Zhang, Ying Jin, Yang Zhang, Zhaojun Liu, Yue Guo, Jingping Yang, Zhihong Liu

## Abstract

Interpreting non-coding variant effects in their native cellular context is the central problem in medical genomics. Using paired single-cell ATAC-seq and whole-genome sequencing in human immune cells, we measured allele-specific regulatory effects at 129,241 heterozygous variants across seven cell types. We trained REG-FOCUS, a transferable sequence-and-chromatin framework, and extended predictions to 1,188,417 common variants across twelve immune cell types. These predictions revealed that enhancer-centric variant directionality is governed by quantitative tuning of transcription factor binding affinity. Applied to systemic lupus erythematosus, REG-FOCUS prioritized causal regulatory variants selectively converging on pathways of approved therapies, and enabled us to identify rare enhancer variants in patients corroborated by expression outliers in carriers. This work brings cell-type-resolved variant interpretation to non-coding personal genomes.

## Main Text

Translating a personal genome into clinical insight is limited not by the detection of genetic variants, but by their interpretation. The overwhelming majority of disease and trait associated variants fall in non-coding regions and act by perturbing cis-regulatory elements in a cell-type-specific manner (*1–3*). The accurate evaluation of regulatory effects of non-coding variants is a prerequisite for translating genetic discoveries into biological and clinical utility.

Deep learning models have advanced the prediction of functional readouts from DNA sequence (*4–7*). However, these models are trained exclusively on reference genomes and do not observe variants or their functional consequences during training, which limits their ability to explain variation across individuals (*8–10*). This has motivated the development of variant-aware models such as PromoterAI which is trained directly on observed variant effects and established clear gains over reference-only inference (*11*, *12*). Yet current work remains constrained by training data restricted to promoter proximal sequences, episomal reporter activity disconnected from native chromatin, or bulk and cell line readouts that dissolve cell-type specificity. Observed variant effects across regulatory elements genome-wide, measured in native chromatin at the resolution of individual cell types, have remained largely unresolved.

Here we address this gap by quantifying cell-type-specific variant effects through paired single cell ATAC-seq (scATAC-seq) and whole-genome sequencing (WGS) of human immune cells, and training a transferable model, REG-FOCUS (REGulatory effects based on Fusion Of Chromatin and Underlying Sequences), on variant-aware measurement. These data and the resulting model together enable a genome wide atlas of cell-type-resolved variant effects across immune lineages and direct prediction of variant effects in individual genomes. We applied this framework to systemic lupus erythematosus (SLE) to interpret non-coding disease-associated variants across the spectrum from common to rare variants.

## Results

### In vivo measurement of regulatory effects of non-coding variants

To directly measure the regulatory effects of variants in their native cellular context, we developed a workflow leveraging paired scATAC-seq and WGS data from primary immune cells. A key design principle was to maximize the causal clarity of each measurement. We employed stratAS (*13*), a statistical framework that explicitly models allele-specific signal in heterozygous individuals, followed by independent chi-square validation to ensure robust effect variant calls (Fig. 1A, Methods). We performed scATAC-seq and WGS on peripheral blood mononuclear cells (PBMCs) from recruited individuals (table S1). ScATAC-seq yielded 152,215 high-quality cells, classified into 12 distinct cell types, including specialized subsets such as age-associated B cells (ABCs) (Fig. 1B and fig. S1). Using phased WGS data, we identified 6,946,151 heterozygous variants (fig. S2), of which 129,241 were retained for analysis after filtering for sufficient read coverage in both datasets. The cell-type-resolved regulatory effects of these variants were evaluated by statistical analysis of allele-specific read counts across cell types (table S2).

**Fig. 1.**
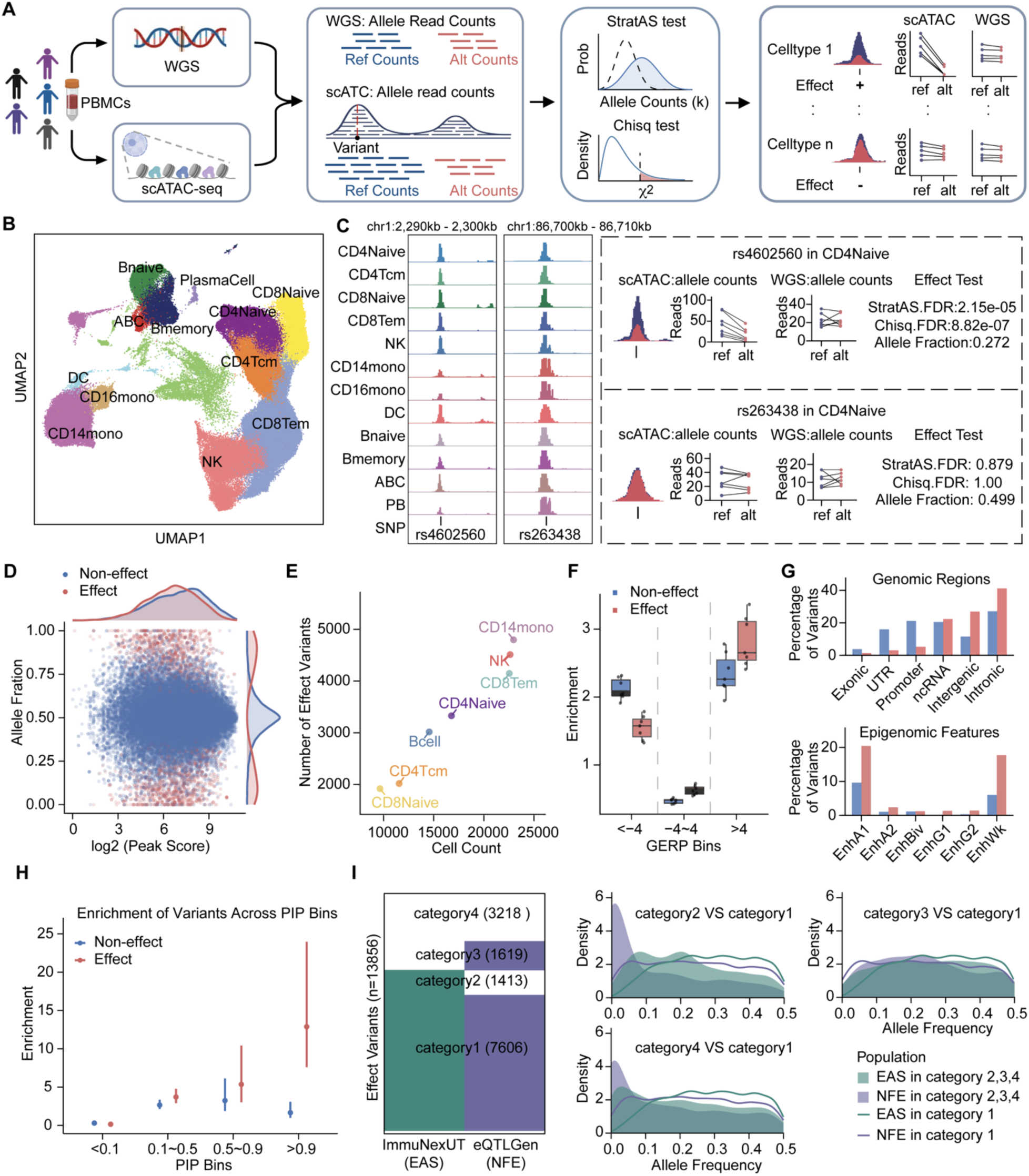
Within-individual allelic imbalance identifies cell-type-resolved regulatory variant effects in primary human immune cells. **(A)** Study design. Paired scATAC-seq and WGS were performed on PBMCs from individuals. Allelic read counts at heterozygous sites from scATAC-seq in each cell type or WGS was quantified in individuals. Variants with significant allelic imbalance passing the stratAS and chi-square test (FDR < 0.05) were defined as effect variants. **(B)** UMAP projection of high-quality scATAC-seq cells, colored by annotated immune cell types. **(C)** Representation of effect and non-effect variants. Genome browser view shows scATAC-seq signal tracks, read pileups, and allele-specific scATAC-seq and WGS reads in heterozygous individuals. Allele fraction = alt reads / (ref reads + alt reads)). **(D)** Scatter plot of peak score and allele fraction for all tested heterozygous variants. Peak score was from MACS2. **(E)** Number of identified effect variants per immune cell type alongside the number of cells used for allele-specific chromatin accessibility analysis. **(F)** Boxplots show the enrichment of variants stratified by GERP score bins. Each data point represents one cell type. **(G)** Bar plots show the proportions of effect (red) or non-effect (blue) variants across genomic regions (top) and ChromHMM-defined chromatin states (bottom). **(H)** Enrichment of effect or non-effect variants on fine-mapped whole-blood eQTLs stratified by posterior inclusion probability (PIP) bins. Error bars indicates 95% confidence intervals. **(I)** Categories of effect variants by overlapping with East Asian (EAS) and/or European (NFE) eQTLs (Left). Alternative allele frequency distributions of variants in category 2, 3, and 4 compared to that in category 1 (Right).

We found that only a small fraction of variants exhibited significant regulatory effects, whereas the majority showed no detectable impact on chromatin accessibility (fig. S3A). For example, rs4602560, a variant located within a region accessible in all cell types, showed significant change on accessibility in CD4 naive and CD4 T central memory cells (Fig. 1C and fig. S3B). In contrast, rs263438, despite residing in accessible region, showed no detectable effects in any cell type (Fig. 1C and fig. S3C). Notably, total read coverage was indistinguishable between effect and non-effect variants, decoupling true biological effects from sequencing depth artifacts (Fig. 1D). Overall, we identified 1,921 to 4,797 variants with significant regulatory effects across seven major cell types, each comprising approximately 10,000 to 25,000 cells (Fig. 1E). The number of detected effect variants correlated positively with cell sample size, reflecting that deeper sequencing depth provides higher statistical power to resolve allelic effects.

These effect variants displayed distinctive genomic characteristics such as significant enrichment in evolutionarily conserved regions (Fig. 1F), and marked preferential localization to distal enhancer elements rather than promoter regions (Fig. 1G). Integration with fine-mapped eQTL datasets showed significantly stronger enrichment for putative causal eQTL variants, with enrichment increasing with posterior inclusion probability (Fig. 1H). These observations indicate that effect variants represent a selectively constrained, enhancer-centric class of regulatory variants.

A critical advantage of individual-level effects measurement over population-level QTL approaches is its power to detect effects of rare variants. We found that 10.20%, 11.68%, and 23.22% of our discovered effect variants were undetected by eQTL analyses in Europeans (*14*), East Asians (*15*), or both populations, respectively (Fig. 1I). These variants had overall lower allele frequencies in the specific population (Fig. 1I), indicating their absence from population studies reflects statistical power limitations inherent to the QTL design rather than lack of functional alteration. Together, these results establish that within-individual allelic effects analysis in paired scATAC-seq and WGS data yields in vivo, cell-type-resolved, regulatory effects of variants spanning the full allele-frequency spectrum, from common to rare.

### Cellular and sequence dependence of regulatory effects

Given the authenticity and highly selective nature of our identified effect variants, we next sought to decipher their molecular characteristics, aiming to uncover the general principles governing regulatory effects. We examined the cell-type specificity of the effect variants and found that the majority were detected in a single cell type (fig. S4A). Storey’s π1 analysis further revealed limited sharing of effect variants between cell lineages, with approximately 10% to 25% overlap among T cells, B cells, and monocytes (Fig. 2A). Even among closely related T cell subtypes, the proportion of shared effect variants was below 75%. Consistent with these observations, allele fractions of effect variants correlated more strongly with eQTL effect sizes in cell-type-matched settings (Fig. 2B). The allele fractions of effect variants from CD14 monocytes exhibited nearly two times higher correlation with eQTL from classical monocytes than that from other cell types. These observations underscored the critical role of cellular context in shaping regulatory effects.

**Fig. 2.**
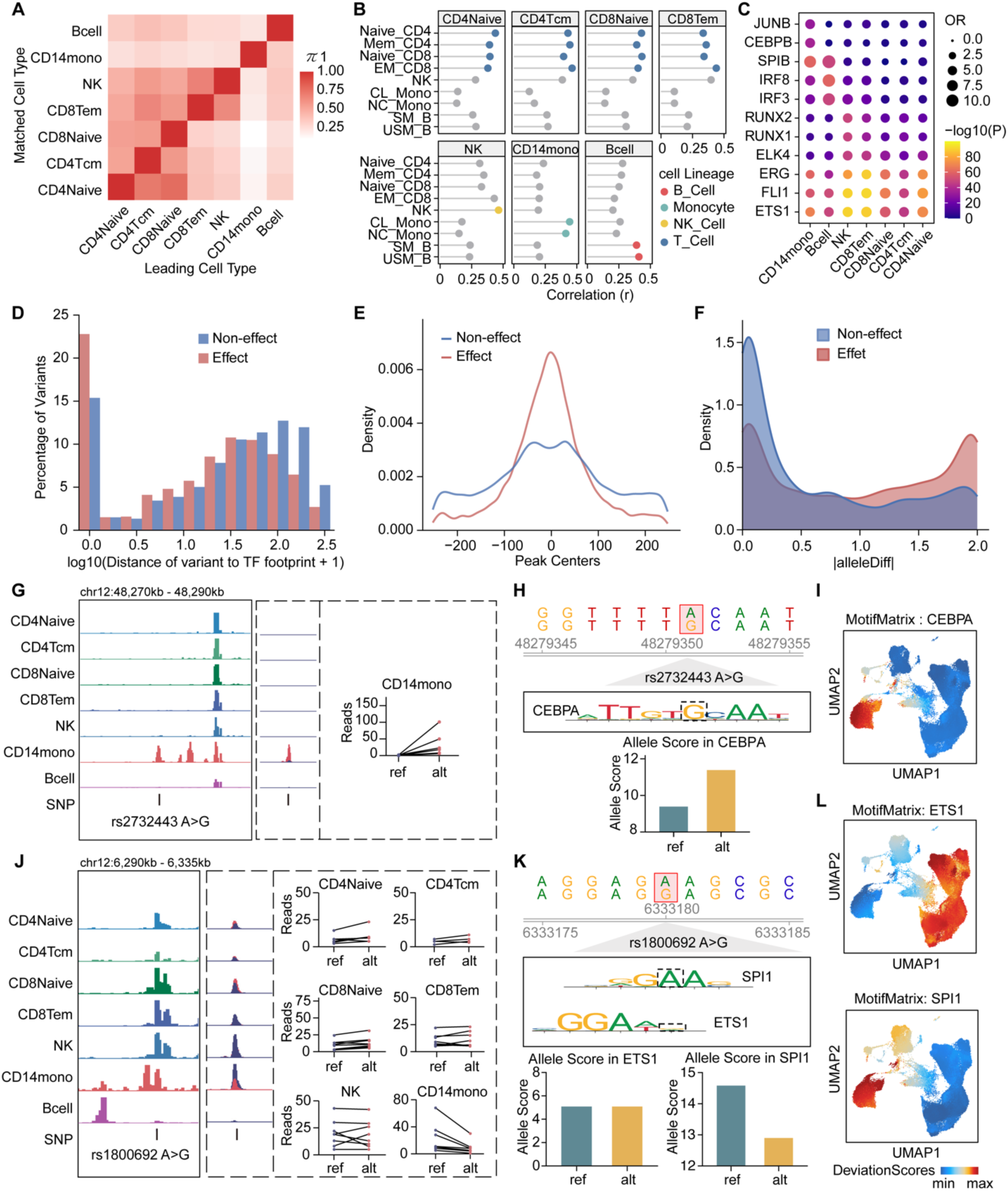
Cell type and sequence context jointly determine the regulatory effects of variants. **(A)** Heatmap of Storey’s π1 statistics for effect variants between cell types. **(B)** Pearson correlation between allele fractions of effect variants in each cell linage and the effect sizes of the variants in eQTLs from ImmuNexUT. **(C)** Motif enrichment of effect variants in each cell type on cell-type-specific TFs. **(D)** Distance distributions from variants to the nearest TF footprint. **(E)** Distance distributions from variants to peak centers. **(F)** TF-DNA binding disruption scores computed using MotifbreakR for variants residing within TF footprints. **(G-I)** Representation of effect variant rs2732443. Genome browser view showing scATAC-seq signal tracks, read pileups, and allele-specific scATAC-seq reads in heterozygous individuals (G). DNA sequence, motif and disruption score by MotifbreakR (H). chromVAR-inferred TF activity for CEBPA (I). **(J-L)** Representation of rs1800692. Genome browser view as G (J). TF binding changes at rs1800692 (K). chromVAR-inferred TF activity for ETS1 and SPI1 (L).

This cellular specificity paralleled transcription factor (TF) targeting. The effect variants from each cell type were preferentially within motifs of their lineage-defining TFs, such as CEBPB motif in monocytes (Fig. 2C and fig. S1D). Footprinting analysis revealed that effect variants were more frequently located within 200 bp of TF binding sites than non-effect variants (Fig. 2D). At the chromatin level, these effect variants were preferentially localized near the centers of accessible regions where the TFs bind (Fig. 2E). Furthermore, MotifbreakR analysis demonstrated that effect variants disrupt TF binding affinity to a greater extent than non-effect variants (Fig. 2F). These results indicate that a variant’s capacity to modulate regulatory effects is strongly dependent on its sequence context, particularly its proximity to and impact on TF binding.

Collectively, these observations suggest that the regulatory effects of variants depend on both cellular and sequence context. Specifically, variants can modulate TF binding by creating or disrupting sequence motifs, but their functional consequences manifest only in cell types where the corresponding TFs are active. For example, variant rs2732443 (A>G) increased chromatin accessibility and regulatory function specifically in monocytes (Fig. 2G and fig. S4B). Motif analysis revealed that this substitution creates a CEBPA binding site (Fig. 2H), a transcription factor with cell-type-specific regulatory activity in monocytes (Fig. 2I). This suggests that the gain of a functional motif leads to increased regulatory activity, but only in the cellular context where the TF is present and active. Conversely, we observed a loss-of-function scenario for rs1800692. Although located in a region accessible in both monocytes and T cells, the A>G substitution selectively reduced accessibility in monocytes but not in T cells (Fig. 2J). Motif analysis showed that this variant disrupts a SPI1 binding site while leaving a nearby ETS1 motif intact (Fig. 2K). Given that SPI1 is active in monocytes whereas ETS1 is active in T cells (Fig. 2L), rs180069 specifically impairs regulatory activity in monocytes (Fig. 2J and fig. S4C), leading to a cell-type-specific loss of accessibility.

Together, these results illustrate a common mechanism whereby the regulatory effects of non-coding variants arises from the interplay between sequence perturbations and cell-type-specific TF activity.

### REG-FOCUS: a modular framework for predicting cell-type-specific regulatory variant effects

Our data revealed that the statistical power for detecting variant effects is constrained by sequencing depth, limiting reliable evaluation to a subset of variants and cell types (Fig. 1E). To overcome this limitation and enable genome-wide prediction, we developed REG-FOCUS, a framework that learns generalizable regulatory logic from measured variant effects and extends predictions to variants and cell types lacking direct effect measurement (Fig. 3A).

**Fig. 3.**
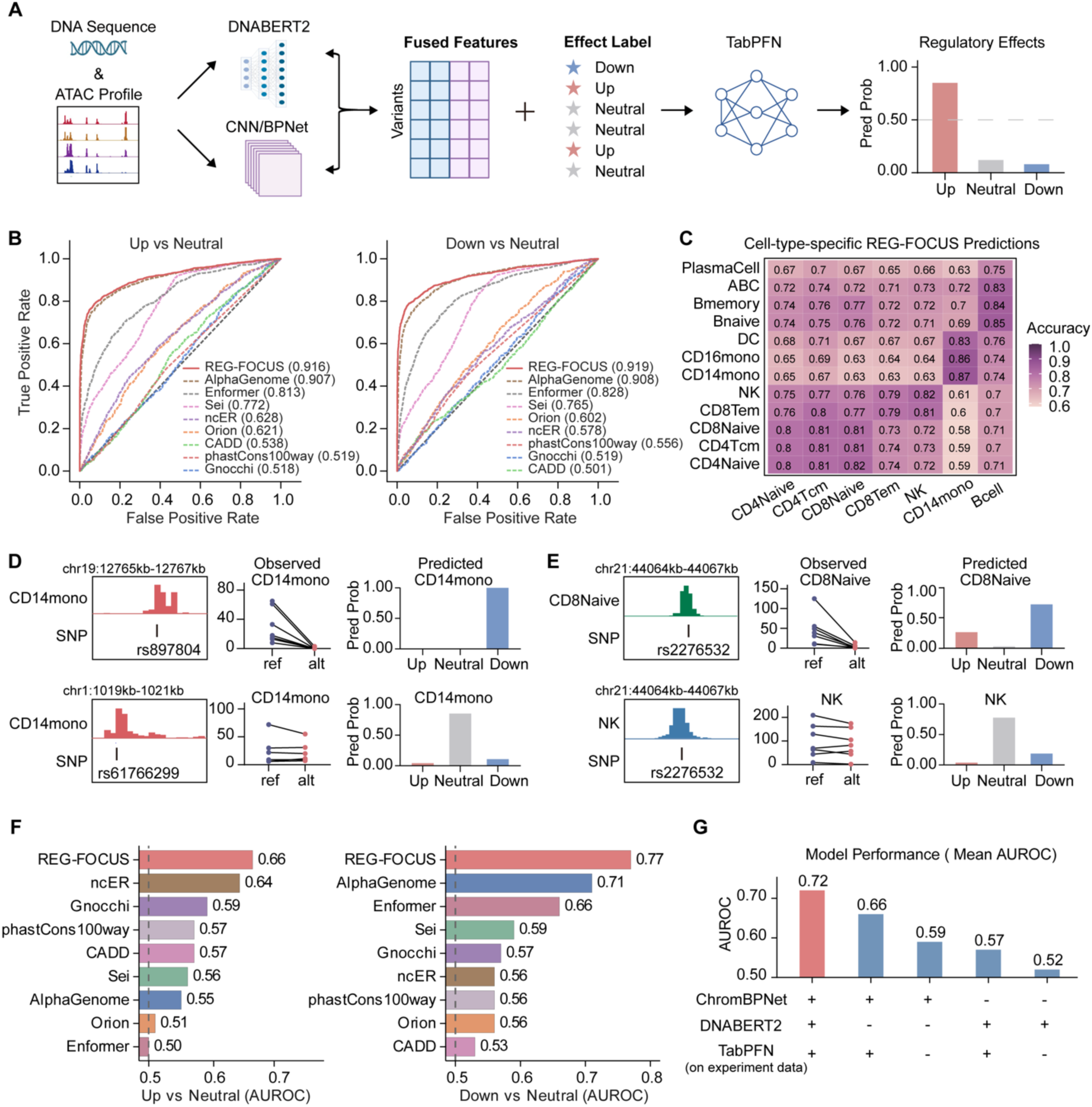
REG-FOCUS accurately predicts regulatory effects across cell types. **(A)** REG-FOCUS model architecture. DNA foundation model (DNABERT2) and chromatin specialized model (ChromBPNet) are fine-tuned and provide complementary feature representations. The joint features are supervised with experimentally derived variant effect labels (Up/Down/Neutral) via TabPFN, establishing a transferable model to directional prediction of regulatory effects across cell types where ATAC profiles are available. **(B)** Predictive performance of REG-FOCUS, benchmark models or scores for groups of variants across six cell types not exposed during training. AUROC values are shown. **(C)** Heatmap shows prediction accuracy when features from each row cell type are fed into REG-FOCUS and effect labels are observed in each column cell type. **(D, E)** Chromatin accessibility track (Left), scATAC-seq reads in heterozygous individuals (Middle), and REG-FOCUS predictions (Right). **(F)** Predictive performance of REG-FOCUS and alternative models or scores on an independent MPRA data. **(G)** Mean AUROC of Up vs. Neutral and Down vs. Neutral predictions was evaluated upon full framework, removal of DNABERT2 features, ChromBPNet features, or TabPFN module on experimentally measured effect labels from training data.

Given that regulatory effects emerge from the interplay between DNA sequence and cellular context, REG-FOCUS integrates two complementary representations, a DNA sequence foundation model (DNABERT2 (*16*)) that captures sequence grammar, and a chromatin-specialized model (ChromBPNet (*17*)) that encodes cell-type-specific chromatin accessibility landscapes. These representations are fused into context-aware features that jointly model intrinsic sequence potential and extrinsic regulatory environment (Table S3). For training data, we restricted variants to those located within open chromatin windows harboring exactly one heterozygous site, thereby ensuring that observed allelic effects can be unambiguously attributed to the single variant present and maximizing the causal clarity of training labels. We characterized regulatory effect directionality by classifying variants as Up, Down, or Neutral based on allele fractions. REG-FOCUS introduces direct effect supervision by coupling these features with experimentally measured effect labels through TabPFN (*18*), a meta-learned tabular reasoning model. Thus REG-FOCUS established a transferable feature-to-effect mapping applicable across cell types which lack effect measurement.

To systematically evaluate the performance of REG-FOCUS, we examined its performance across multiple cell types with experimentally measured regulatory effects. On held-out variants within CD14 monocytes (the training cell type), REG-FOCUS achieved AUROC (area under the receiver operating characteristic curve) over 0.9 for all pairwise comparisons among Up, Down, and Neutral categories (fig. S5A). Critically, when applied to other six cell types where no variant effect measurement were provided during training, the model maintained high performance, with both AUROC and AUPRC (area under the precision-recall curve) stably around 0.9 (Fig. 3B and fig. S5B-C). Furthermore, the predictions showed higher accuracy when the target cell types were matched (Fig. 3C). It can accurately distinguish the effect and non-effect variants in a cell-type-specific manner. For example, the predictions in the CD14 monocyte clearly inferred the down-regulating effect of rs897804 and the neutral variant rs61766299, in agreement with experimental observations (Fig. 3D). REG-FOCUS also successfully recapitulated the down-regulating effect of rs2276532 in CD8 naive T cells but not in NK cells (Fig. 3E). We compared the performance of REG-FOCUS with a range of existing variant effect prediction methods, including deep learning models (AlphaGenome (*6*), Enformer (*4*), and Sei (*5*)) as well as widely used pathogenicity and constraint-based scores (ncER (*19*), Orion (*20*), CADD (*21*), PhastCons100way (*22*), and Gnocchi (*23*)). The results showed that REG-FOCUS achieved the highest predictive performance among all evaluated methods (Fig. 3B and table S4). These results indicate that REG-FOCUS successfully learns a transferable mapping from sequence and chromatin context to regulatory effects.

When we applied REG-FOCUS on a MPRA dataset (*24*), an independent type of experimental assessment of variants effects, the result showed that REG-FOCUS consistently performed best across the evaluation metrics (Fig. 3F and table S5). To further dissect the contribution of individual components within REG-FOCUS, we performed ablation analyses on the MPRA dataset. Compared to the full model, ablation of any module reduced the performance. Notably, the performance degradation further when experimental data were excluded. These results indicate that the availability of real functional consequence dataset is critical for the predictive performance of REG-FOCUS (Fig. 3G). Together, these results demonstrate that REG-FOCUS not only improves predictive accuracy, but more importantly, enables transferable, cross-context inference of regulatory variant effects.

### A regulatory variant atlas reveals enhancer-centric logic in immune cells

Leveraging REG-FOCUS, we systematically evaluated the regulatory effects of variants across the genome with cell-type-specificity, constructing a cell-type-resolved atlas for 1,188,417 common variants. This resource spans to twelve immune cell types, including rare populations such as ABCs (Fig. 4A and table S6). To experimentally validate the predictions, we performed allele-specific luciferase reporter assays on selected variants. Consistent with our predictions, the variants identified as down-regulating effects exhibited significant allele-specific reduction in reporter activity, while those predicted to be neutral displayed comparable activity between alleles (Fig. 4B and fig. S6). Across cell types, approximately 300,000 to 400,000 SNPs resided within accessible regions. Of these, only 9% were predicted to exert regulatory effects in at least one cell type. Up-regulating and down-regulating variants accounted for about 3.5% and 5.2% of these accessible variants, respectively (Fig. 4A). These results indicate that the majority of variants within accessible chromatin do not alter regulatory activity, underscoring the necessity of effect-based variant prioritization beyond positional annotation alone.

**Fig. 4.**
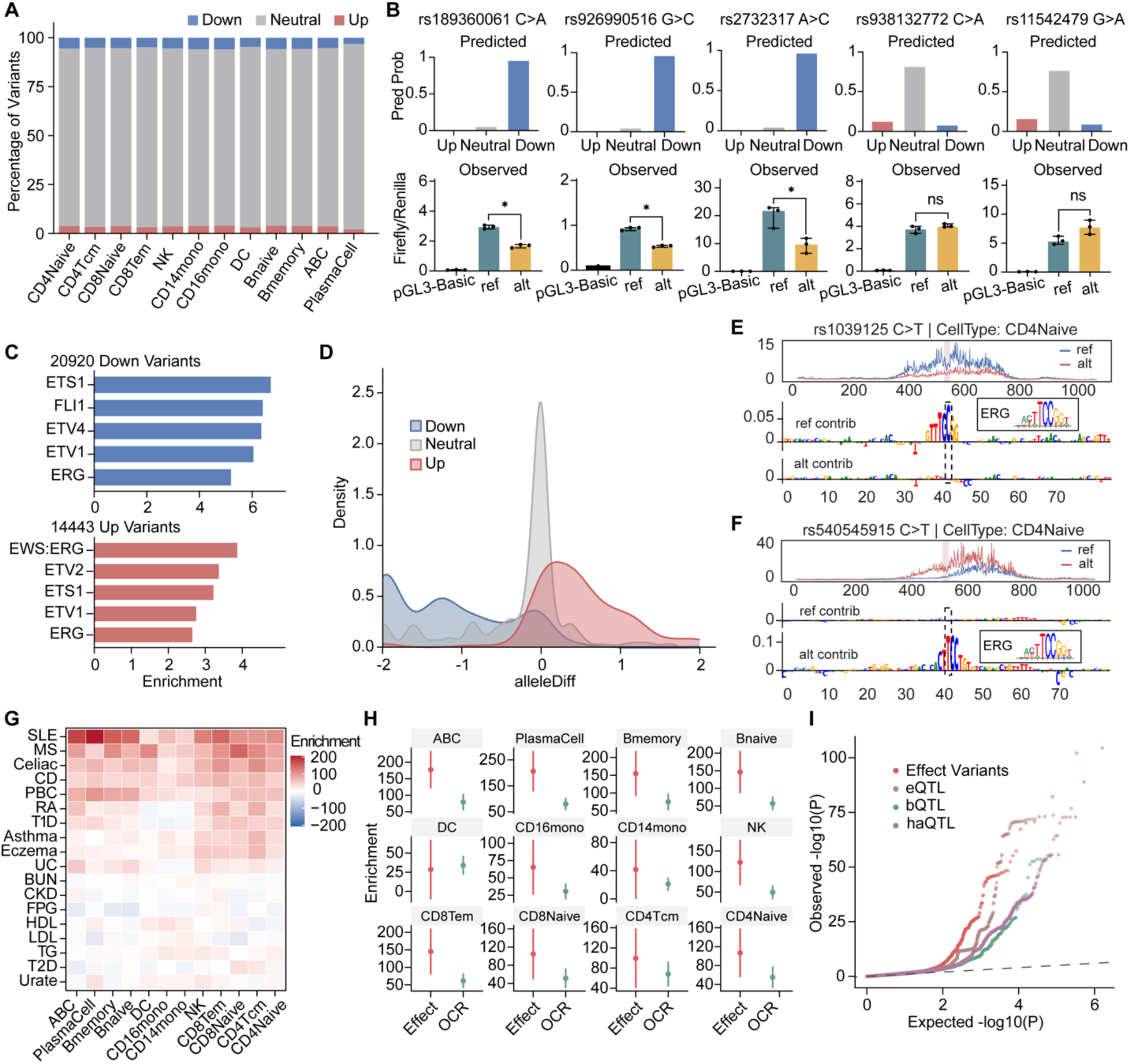
A genome-wide atlas of cell-type-resolved regulatory variant effects. **(A)** Proportions of predicted effect variants (Up/Down) and non-effect variants (Neutral) across 12 immune cell types. **(B)** Validation of predicted effects of variants by allele-specific luciferase reporter assay. Upper panels show REG-FOCUS predictions. Lower panels show results from luciferase reporter assays (n = 3). * represents P < 0.05. **(C)** The top five enriched TF motifs for up and down variants in CD4 naive T cells **(D)** TF-DNA binding affinity changes of up, down, and neutral variants for the top five TF motifs identified in (c), computed using MotifbreakR. **(E, F)** Predicted chromatin accessibility (top) and sequence contribution scores (bottom) at rs1039125 (E) and rs540545915 (F) show effects on the same motif. **(G)** Heatmap shows LDSC-based partitioned heritability enrichment coefficients for ±50-bp windows around effect variants across immune cell types for immune and control traits. SLE, systemic lupus erythematosus; MS, multiple sclerosis; CD, crohn’s disease; PBC, primary biliary cirrhosis; RA, rheumatoid arthritis; T1D, type 1 diabetes; UC, ulcerative colitis; BUN, blood urea nitrogen; CKD, chronic kidney disease; FPG, fasting plasma glucose; HDL, high-density lipoprotein; LDL, low-density lipoprotein; TG, triglycerides; T2D, type 2 diabetes. **(H)** Heritability enrichment of effect variant regions (±50 bp) or all open chromatin regions (OCR) for SLE across cell types. **(I)** Quantile-quantile (QQ) plots of SLE GWAS association P values for variants annotated as predicted effect variants, eQTLs, haQTLs, and bQTLs.

Consistent with experimentally identified effect variants (Fig. 1G), both predicted up- and down-regulating variants were predominantly localized to enhancer regions (fig. S7), highlighting an enhancer-centric distribution of regulatory variants. Previous studies of promoter variants have suggested that regulatory directionality is largely determined by the functional class of the disrupted TF (activators versus repressors) (*12*). Whether this paradigm extends to the enhancer context is not clear. We found that genome-wide effect variants are preferentially enriched in cell-type-specific TF motifs (fig. S8), consistent with their functional relevance. However, in contrast to promoter-centric variants, the genome-wide variants predicted to exert opposite regulatory effects were enriched for same TF motif classes within the cell type. For example, in CD4 naive T cells, both up- and down-regulating variants showed strong enrichment for ETS family motifs (Fig. 4C), indicating that TF identity alone does not determine regulatory directionality in the enhancer context. Instead, analysis of TF binding affinity revealed a clear divergence. The up-regulating variants tended to enhance TF-DNA binding, whereas down-regulating variants preferentially disrupted binding (Fig. 4D-F). SNP-SELEX-based affinity measurement (*25*) showed the same trend between predicted binding changes and directional variant effects (fig. S9). These results suggest that, at enhancer-centric variants, regulatory directionality is closely associated with quantitative modulation of TF binding affinity within a shared TF, a principle distinct from the TF-class-based model proposed for promoter variants.

We further investigated the contribution of these effect variants to diseases. Partitioned heritability analysis revealed that effect variants contributed substantially more to the heritability of immune diseases than to non-immune traits, with highly cell-type-specific enrichment patterns (Fig. 4G). For SLE, effect-variants-derived heritability was most strongly enriched in B cell subsets such as ABCs, consistent with the central role of B cells in lupus pathogenesis (*26*, *27*). For rheumatoid arthritis (RA), enrichment was pronounced in T lymphocytes, especially CD4 T cell subsets (Fig. 4G), in agreement with the established role of T cells in RA pathophysiology (*28*). Importantly, within individual diseases such as SLE, genomic regions harboring effect variants showed significantly greater heritability enrichment compared to overall accessible chromatin regions (Fig. 4H), indicating that effect variants capture functional signals beyond general chromatin accessible regions. Furthermore, relative to variants derived from other functional genomic datasets such as eQTLs, haQTLs (*29*), bQTLs (*30*), our effect variants exhibited a stronger deviation from the null distribution in QQ plots (Fig. 4I), suggesting an increased likelihood of causal involvement in disease-associated regulatory perturbations.

### Regulatory effect-based prioritization of SLE causal variants

To directly infer functional causality, we applied a one-step in silico screening to prioritize GWAS variants based on their predicted regulatory effects. For all variants in linkage disequilibrium (LD) with lead SNPs, we applied REG-FOCUS to quantify their regulatory effects across immune cell types, and prioritized effect variants as putative causal ones (Fig. 5A).

**Fig. 5.**
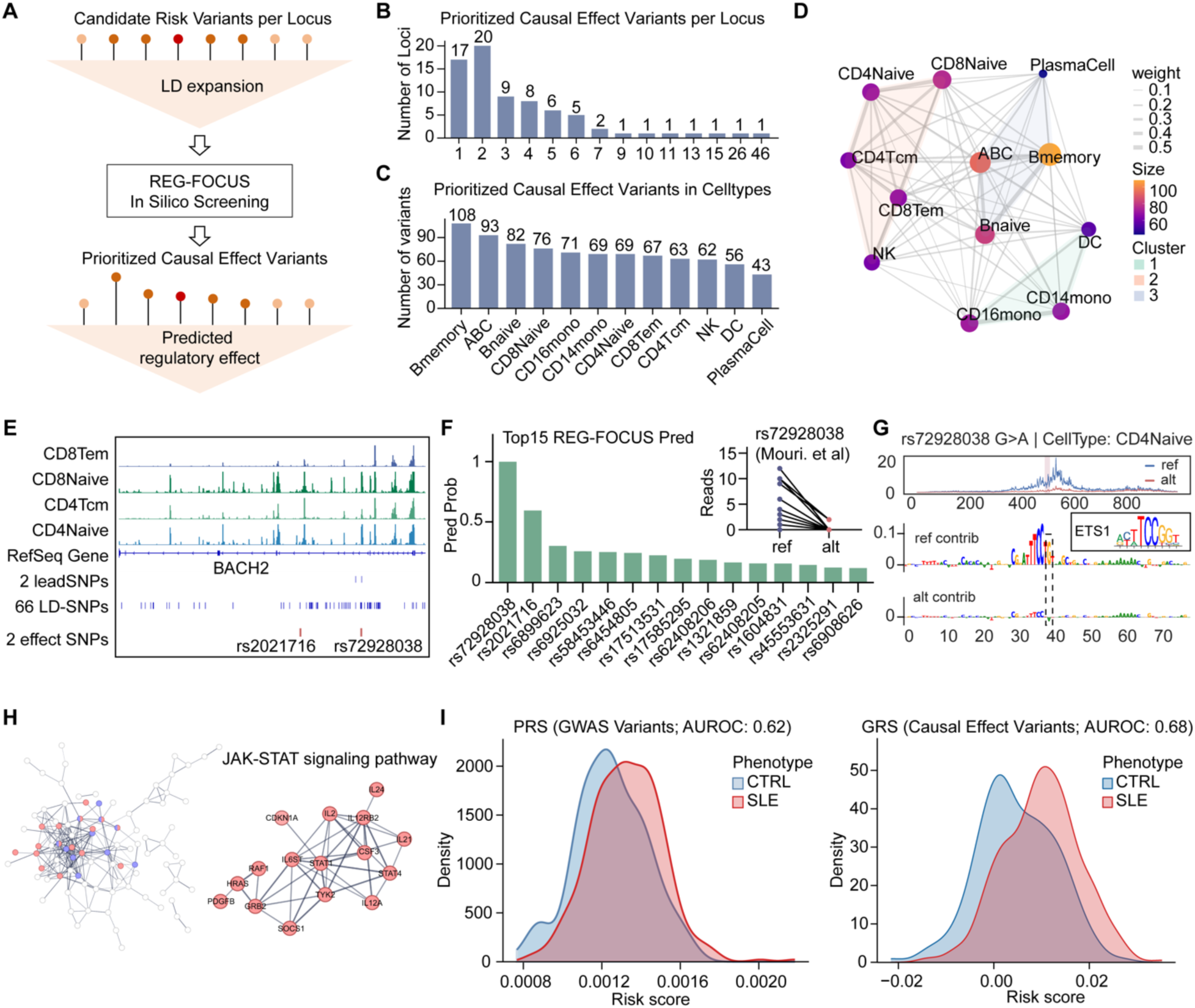
Effect prioritization refines causal variant mapping at SLE GWAS loci and improves genetic risk stratification. **(A)** Schematic overview of the effect-guided one step in-silico screening strategy for prioritizing GWAS causal variants. **(B)** Distribution of identified causal variant counts per locus (n = 74). The x-axis indicates the number of causal variants per locus, and the y-axis shows the number of loci. **(C)** Distribution of causal variants across immune cell types. **(D)** Cell-type sharing network of causal variants. Node size and color reflect the number of causal variants identified in each cell type. Edge width is proportional to the sharing fraction. Network communities were identified using the Walktrap algorithm. **(E-G)** Representation of variants screening at BACH2 locus. Tracks for chromatin accessibility, lead SNPs, SNPs in LD blocks with lead SNPs, and prioritized effect SNPs (E). The top 15 predictions in CD4 naive T cells, and allele-specific reads for top SNP rs72928038 in the insert (F). Predicted effects and sequence contribution of rs72928038 (G). **(H)** Protein-protein interaction (PPI) network of target genes linked to identified causal variants. The full network representation (Left) and the top enriched JAK-STAT module (Right). **(I)** Polygenic risk score (PRS) modeled from all GWAS variant (Left) and genetic risk score (GRS) derived from prioritized causal variants (Right) for SLE patients and controls (n = 311 SLE patients and 178 healthy controls).

Applying this approach to a large-scale SLE GWAS dataset (*31*), we mapped 15,910 risk variants to 320 putative causal variants across 74 risk loci (Fig. 5B and table S8). Lead SNPs in the GWAS had only a 3% probability of representing a causal variant. While B cells showed the strongest enrichment of candidate variants, consistent with their established role in SLE, other cell types also made substantial contributions (Fig. 5C), revealed a multi-cellular genetic architecture underlying disease susceptibility (Fig. 5D).

This framework not only refines causal variant within LD blocks, but also enables mechanistic interpretation. At the well-studied BACH2 locus, we narrowed 66 LD variants to two effect SNPs (Fig. 5E). Among them, rs72928038, predicted to have the strongest regulatory effect (Fig. 5F). rs72928038 has been previously validated as a causal variant (*32*), serving as a positive control. The prediction not only showed that G>A substitution markedly altered chromatin accessibility but also disrupted a key ETS1 binding motif (Fig. 5G), illustrating a direct sequence-to-function link. Beyond known loci, our approach uncovered previously underappreciated regulatory variants. At a distinct risk locus, rs7769906 was prioritized as the top candidate (fig. S10A). In ABCs, the risk allele C (ref allele) showed lower chromatin accessibility and expression of DEF6 (fig. S10B-D), a critical regulator restraining the differentiation and expansion of ABCs (*33*).

Based on the prioritized variants, we mapped target genes and constructed a protein-protein interaction network. The most prominent functional module was the JAK-STAT signaling pathway (Fig. 5H), a central immune signaling axis implicated in SLE pathogenesis and widely recognized as a therapeutic target (*34*, *35*). This pathway enrichment was specific to genes linked to prioritized variants but absent from that of all GWAS variants, highlighting the ability of effect-based prioritization to uncover biologically coherent disease modules. We then applied the prioritized variants to genetic risk scores (GRS) on our SLE cohort (independent from the discovery GWAS). The GRS based solely on prioritized causal variants significantly outperformed the standard one derived from all GWAS variants (AUROC: 0.68 vs. 0.62), despite using substantially fewer variants (Fig. 5I). This observation underscores that effect-informed prioritization enriches true causal signals, enhancing genetic risk interpretation.

### Extending disease gene discovery to rare non-coding variants

As rare variants contribute substantially to disease, yet their interpretation has been largely restricted to coding regions. The coding variants in known SLE susceptibility genes explain only 4% of patients in SLE cohort (*36*), suggesting non-coding regulatory variants in these genes contribute to the remaining genetic basis. To address this gap, we leveraged REG-FOCUS to predict regulatory effects of rare non-coding variants and analyzed individual WGS data from a cohort of 311 SLE patients and 178 healthy controls. Using effect-informed variants for gene-based association analysis (BeviMed (*37*)), we found that prioritized genes were significantly enriched for known SLE pathogenic genes such as IRF5 and BACH2 (Fig. 6A), demonstrating that regulatory effect-informed rare variants analysis can recover disease genes in the non-coding space.

**Fig. 6.**
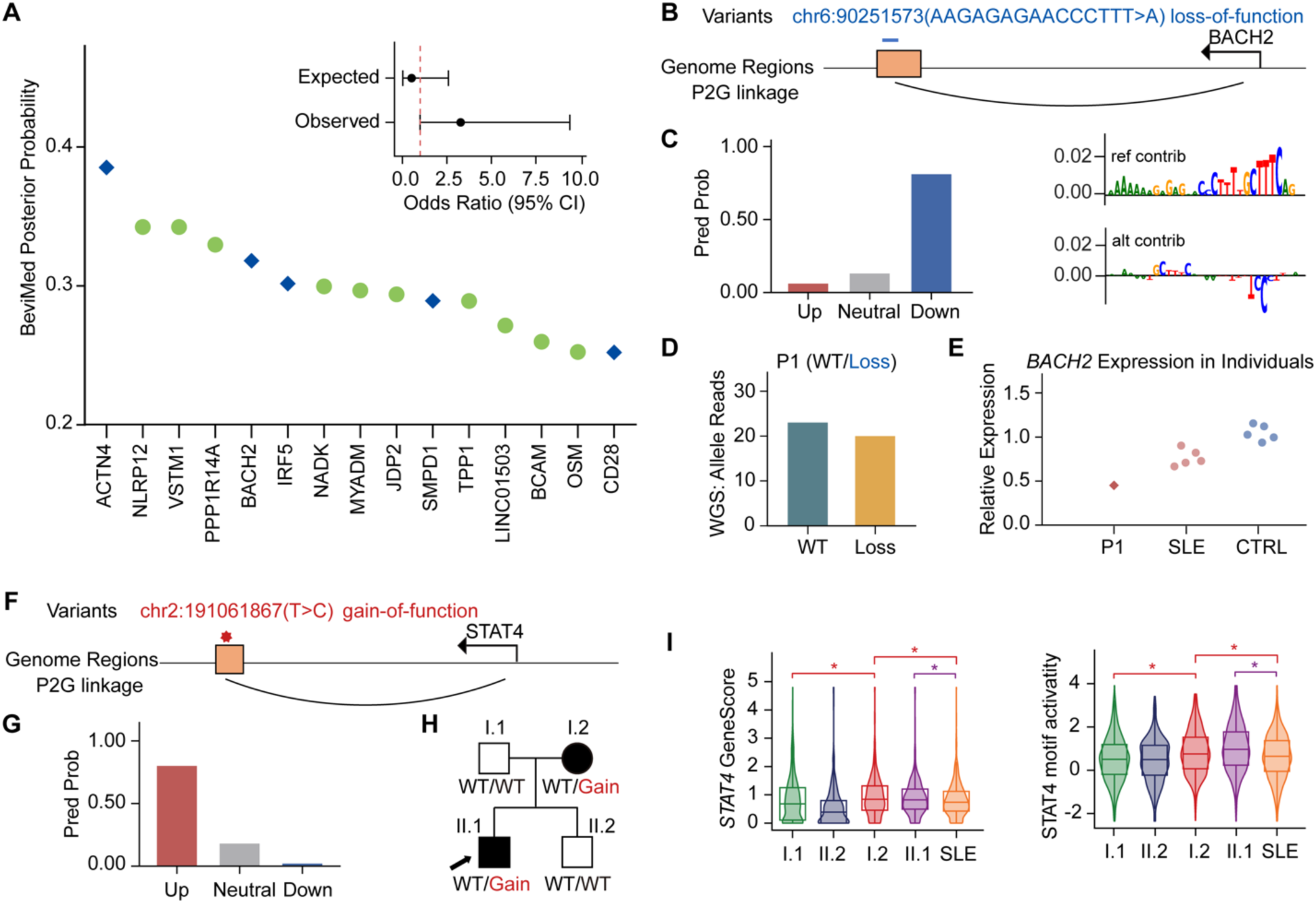
Interpretation of rare non-coding variants in individuals based on regulatory effects. **(A)** Gene-level association analysis based on rare non-coding effect variants using BeviMed. The top 15 genes ranked by posterior probability of association (PPA) are shown. Known SLE genes are highlighted in blue. The inset shows enrichment of known SLE genes among the top 15 genes. **(B-E)** Representation of a novel loss-of-function variant at regulatory region of *BACH2*. Genomic location and peak2gene linkage of the variant (B). REG-FOCUS prediction and sequence contribution of the variant in naive B cells (C). WGS-confirmed carrier P1 of the variant (D). Relative expression of *BACH2* in P1, SLE non-carriers, and healthy controls (E). **(F-I)** Representation of a rare gain-of-function variant at regulatory region of *STAT4*. Genomic location and peak2gene linkage of the variant (F). REG-FOCUS prediction in CD4 naive T cells (G). Pedigree of the family carrying the variant (H). The proband (II.1) is indicated by an arrow. Affected and unaffected individuals are indicated by black and white symbols, respectively. *STAT4* gene activity (left) and chromVAR-inferred STAT4 TF activity (right) in scATAC-seq in CD4 naive T cells in carriers, non-carrier family members and an independent SLE patient. Statistical significance was assessed using a two-sided Mann–Whitney U test (*P < 0.05).

Our framework extends the contribution of established SLE genes beyond coding variants. At the *BACH2* locus, a key regulator of immune tolerance (*38*), we identified a novel regulatory indel chr6:90251573(AAGAGAGAACCCTTT>A) located within regulatory region of *BACH2* (Fig. 6B). REG-FOCUS predicted that the sequence change significantly reduced chromatin accessibility at this site (Fig. 6C). Consistent with this loss-of-function prediction, *BACH2* expression was decreased in the carrier compared to non-carriers as well as healthy controls, confirmed by WGS and expression data (Fig. 6D, E and fig. S11A). We also observed a gain-of-function effect at the regulatory element of *IRF5*, a central regulator of type I interferon signaling (*39*). A rare variant chr7:129072795(T>C) (MAF = 1e-5) was predicted to increase chromatin accessibility, and *IRF5* expression was significantly elevated in the carrier compared to both non-carrier SLE patients and healthy controls (fig. S11B and fig. S12). These results demonstrate that effect rare non-coding variants can modulate established disease genes significantly.

We further investigated segregating regulatory variants within families. At the *STAT4* locus, one of the strongest genetic risk loci for SLE (*40*), we identified a novel variant chr2:191061867(T>C) located within regulatory region of *STAT4* (Fig. 6F). REG-FOCUS predicted increased regulatory activity at this variant (Fig. 6G). Pedigree analysis confirmed that this variant is shared by two affected family members but absent in unaffected relatives (Fig. 6H and fig. S11C-D). Consistent with the gain-of-function prediction, the single-cell analysis in individuals revealed that not only the *STAT4* expression was elevated, but also the transcription factor activity (inferred by chromVAR) was increased in carriers (Fig. 6I).

Collectively, these findings demonstrate that regulatory effects prediction extends the interpretation of disease genetics from coding to non-coding space, thereby increasing the fraction of patients whose genetics can be functionally interpreted.

## Discussion

Non-coding variants are major contributors to disease etiology, but elucidating the clinical significance remains challenging. In this study, with each individual’s heterozygous sites as in vivo perturbations, we measure the cell-type-resolved regulatory effects of non-coding variants. Training on the variant-aware dataset, REG-FOCUS improves variant effects prediction and generalizes to diverse primary cell types. We yield a comprehensive regulatory effects atlas of common variants across twelve immune cell types. Furthermore, through our effect-variant strategy, we demonstrate the clinical relevance of non-coding variants, from population-scale prioritization of common risk variants to individual-level analysis of rare variants in SLE.

Genomic non-coding elements orchestrate gene expression programs and define cellular identity(*41*, *42*). Their perturbation by non-coding variants contributes to human diseases (*43–46*). Recent PromoterAI has successfully predicted the effects of promoter variants and demonstrated the clinical utility of variant-aware learning (*12*). However, other classes of regulatory elements such as enhancers remain less explored. By measuring the effects of genomic variants, we observed a significant enrichment of effect variants across classes of non-coding elements, particularly within enhancers. Furthermore, we found that genomic variants diverge from promoter variants at TF regulatory logic, consistent with previous findings (*47*, *48*). These observations highlight the necessarily of modeling effects of variants beyond promoter proximal ones. With training on genomic non-coding variants, REG-FOCUS can predict the effects of variants across types of regulatory elements. Our work broadens the regulatory landscape of non-coding variants, and leverage the utility of WGS on understanding of disease etiology.

Non-coding elements exert their roles through cell-type-specific regulatory function. Accurate evaluation of variant effects in their native cellular context is required to capture their cell-type-specific regulatory impact. With the design of modular framework, REG-FOCUS is readily generalizable to diverse cell types where chromatin accessibility profiles are available. It enables the prediction of variants effects in disease-associated cell types, especially those are difficult to isolate or culture, such as ABCs. Recent efforts, such as the Human Dynamic Multi-Organ Atlas (HDMA) (*49*) and Chinese Immune Multi-Omics Atlas (CIMA) (*50*), have generated chromatin accessibility landscapes across various organs and cell types. Combined with these data recourses, REG-FOCUS will extend variant-aware effect inference beyond the immune system and facilitate genetic interpretation of various diseases outside the immune traits. Furthermore, the modular design of REG-FOCUS allows its component such as DNABERT2 and ChromBPNet to be readily replaced with improved foundation or specialized models as they become open-access, enabling iterative enhancement of predictive performance.

Our study evaluated the effects of variants on chromatin accessibility. Although previous studies have shown that variants disrupting accessibility and TF motifs can lead to concordant changes in gene expression (*49*), there is still a gap between the effects on chromatin accessibility and the effects on expression. In this study, we bridged the gap by using the established Peak2Gene linkage. However, to better understand the role of non-coding variants in disease, a more quantitative approach is needed to link their effects on chromatin accessibility to downstream gene expression changes(*51*). Despite this limitation, our work provides a foundation for refining the clinical interpretation of non-coding variants in cell-type-specific manner and leverage the utility of WGS on precision medicine for complex diseases.

## Data Availability

All data produced in the present study are available upon reasonable request to the authors

## Author contributions

**Conceptualization:** Z.H.L., J.P.Y., Q.L.

**Methodology:** J.P.Y., Q.L.

**Investigation:** Q.L., J.Z., Y.P.Z., K.C.W., C.M.Z., J.H.Z., Y.J., Y.Z., Z.J.L., G.Y.

**Visualization:** Q.L., J.P.Y.

**Funding acquisition:** Z.H.L., J.P.Y.

**Project administration:** Z.H.L., J.P.Y.

**Supervision:** Z.H.L., J.P.Y.

**Writing – original draft:** Q.L.

**Writing – review & editing:** Z.H.L., J.P.Y.

## Supplementary Figures

**Fig. S1.**
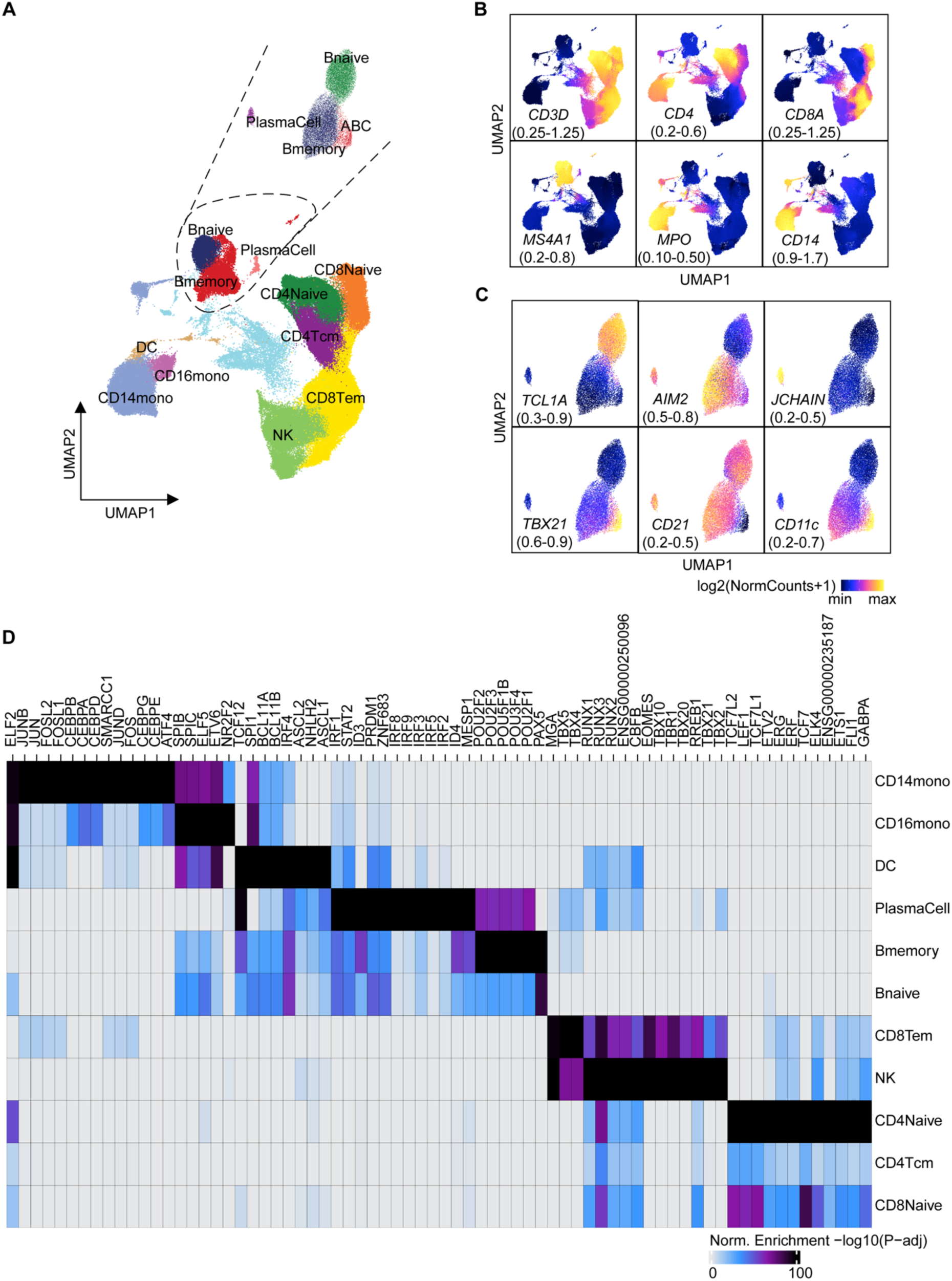
Single-cell chromatin landscape and regulatory programs of human peripheral immune cell types. **(A)** UMAP embedding of PBMCs colored by annotated clusters corresponding to 12 cell types. Inset shows UMAP projection of B cells re-clustered into four subpopulations: naive B cells, memory B cells, plasma cells, and age-associated B cells (ABCs). **(B, C)** Density distributions of gene scores for canonical marker genes across PBMCs (B). Gene scores for canonical B cell marker genes (C). **(D)** Enrichment of TF motifs in marker peaks for each immune cell types.

**Fig. S2.**
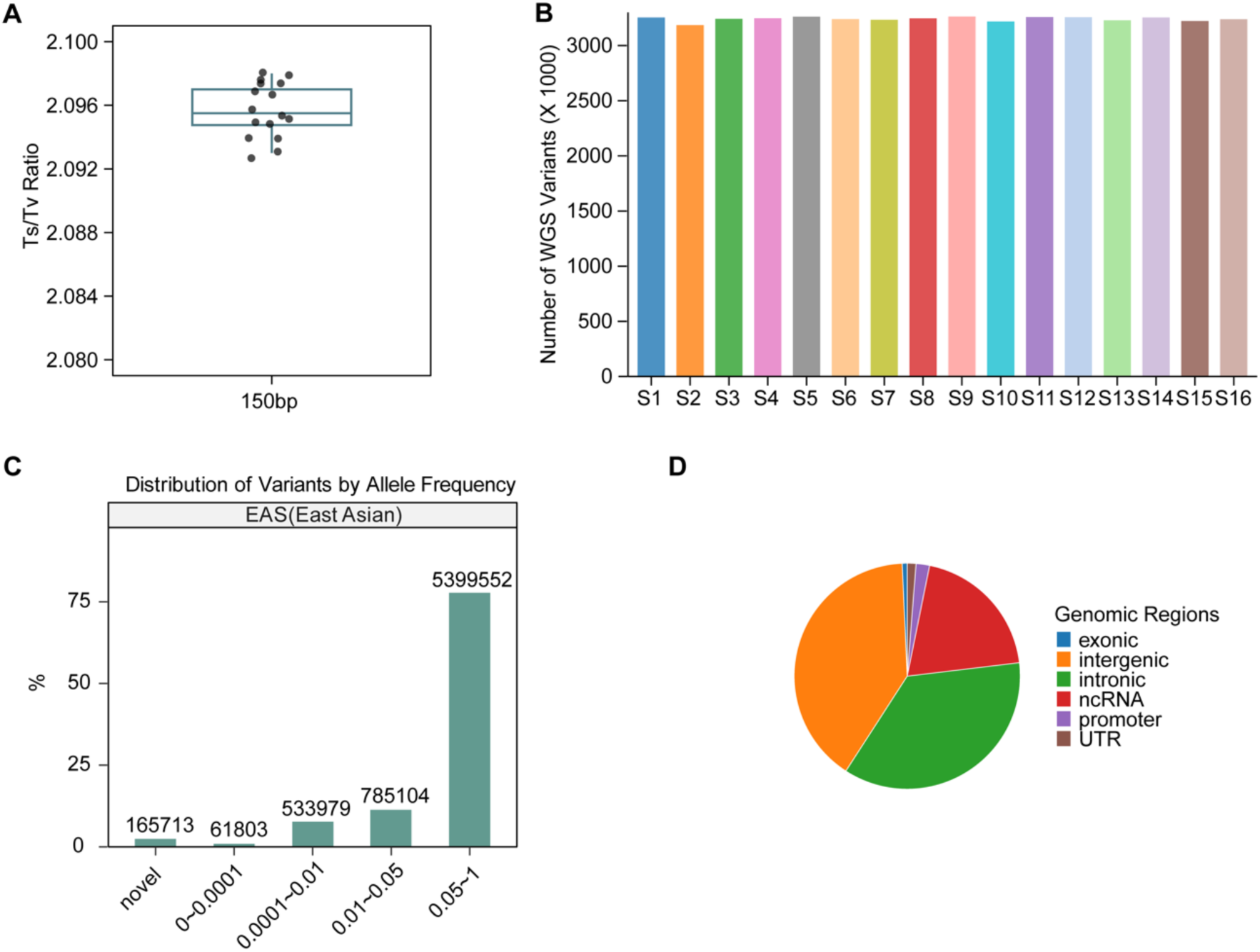
Variants discovery from WGS data in individuals. **(A)** Boxplots showing transition/transversion (Ts/Tv) ratios across WGS samples (n = 16; 150-bp reads). **(B)** Number of variants per sample following stringent quality control. **(C)** Proportional distribution of variants across allele frequency bins in the East Asian population from gnomAD v4.1.0. Novel variants are defined as those absent from gnomAD v4.1.0. **(D)** Distribution of variants across annotated genomic regions.

**Fig. S3.**
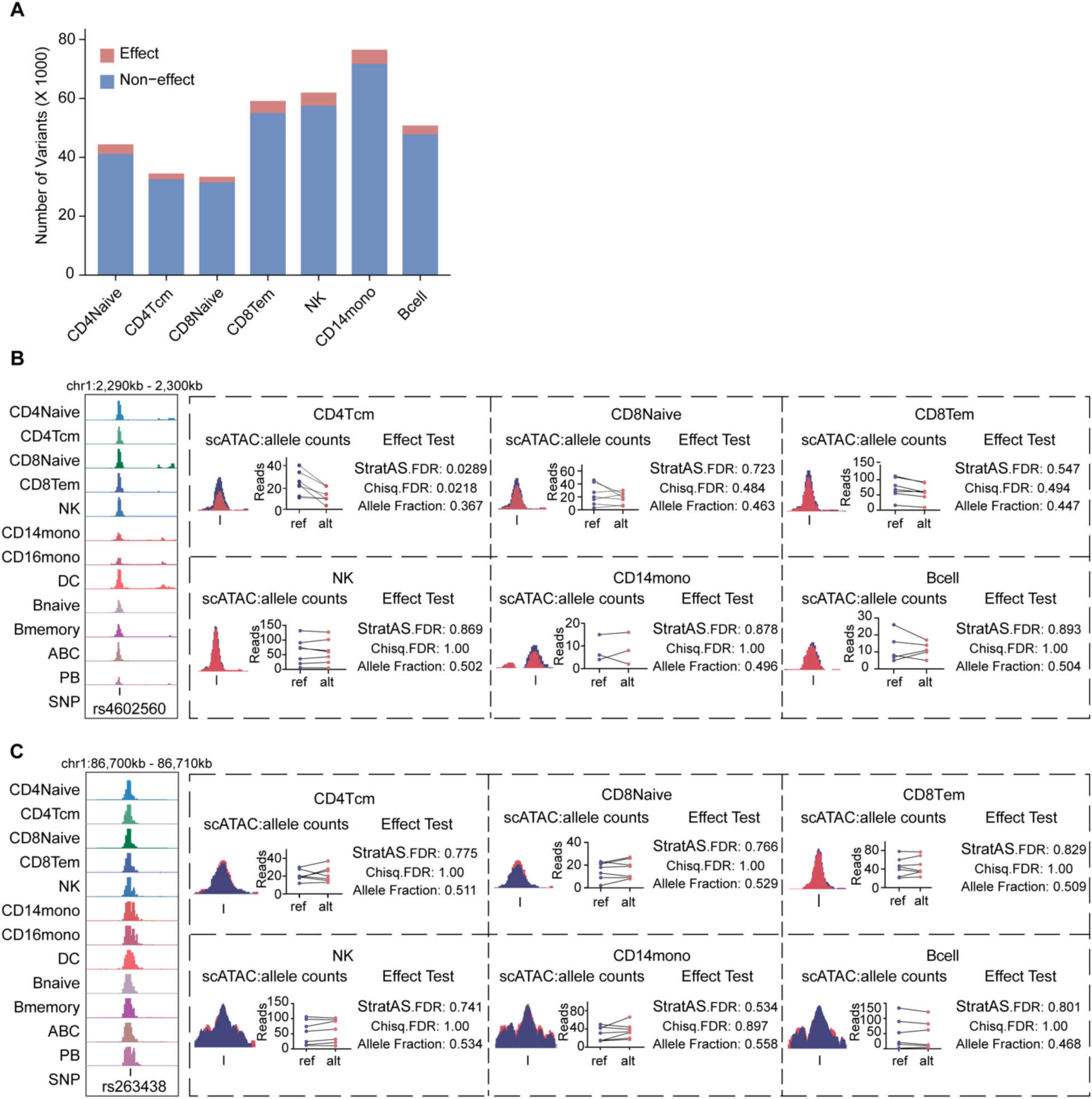
Regulatory effects of non-coding variants on chromatin accessibility across immune cell types. **(A)** Number of variants included in the analysis of allelic effects on chromatin accessibility across seven immune cell types. **(B, C)** Allelic effects of rs4602560 (B) and rs263438 (C) on chromatin accessibility in immune cell types. The detail information described at Fig. 1C.

**Fig. S4.**
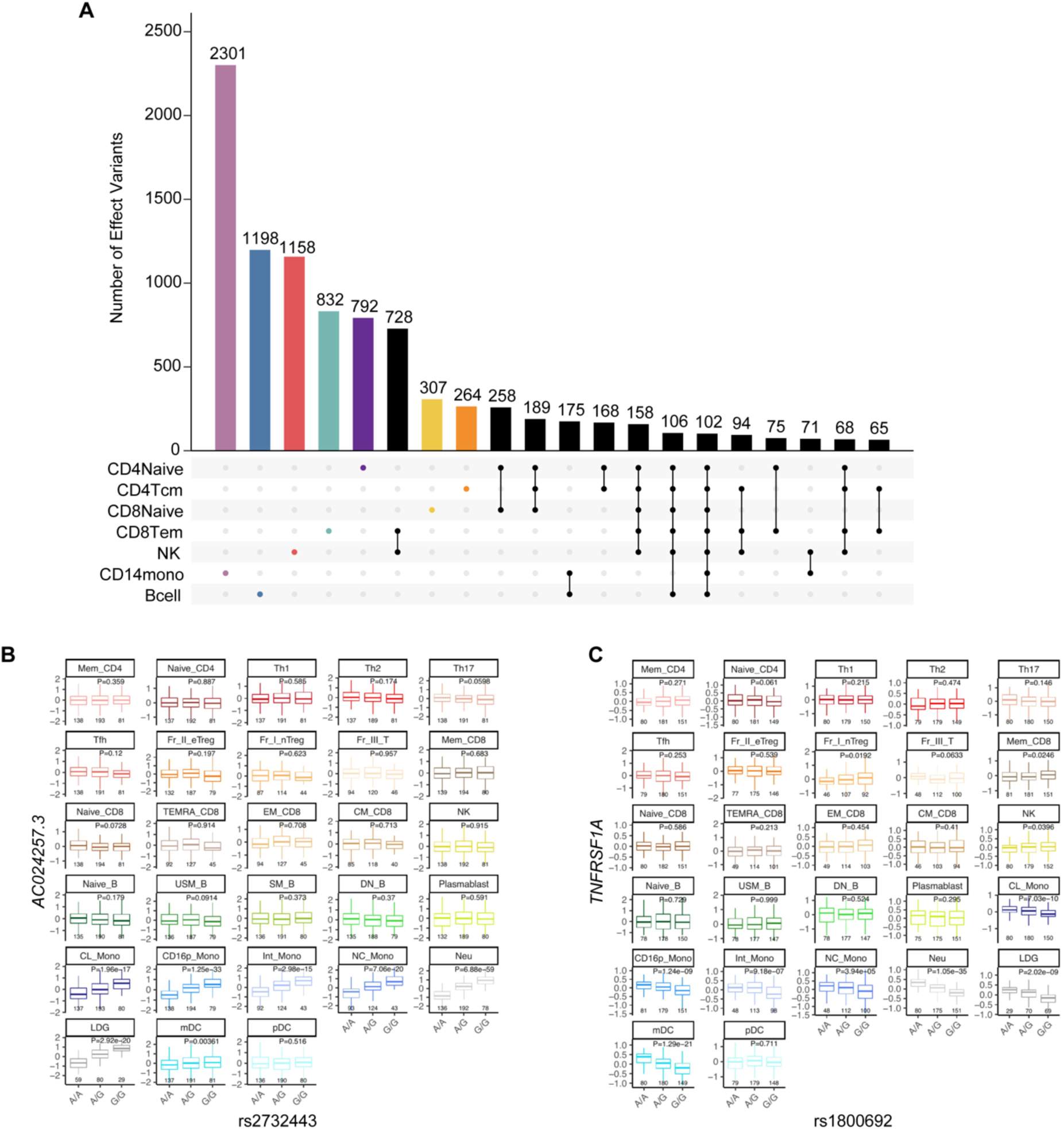
Cell-type-specific regulatory effects of non-coding variants across immune cell types. **(A)** Upset plot of effect variants across immune cell types. Bar plots show the size of intersecting sets. **(B, C)** eQTL associations for rs2732443 (B) and rs1800692 (C) across immune cell types are derived from the ImmuNexUT dataset.

**Fig. S5.**
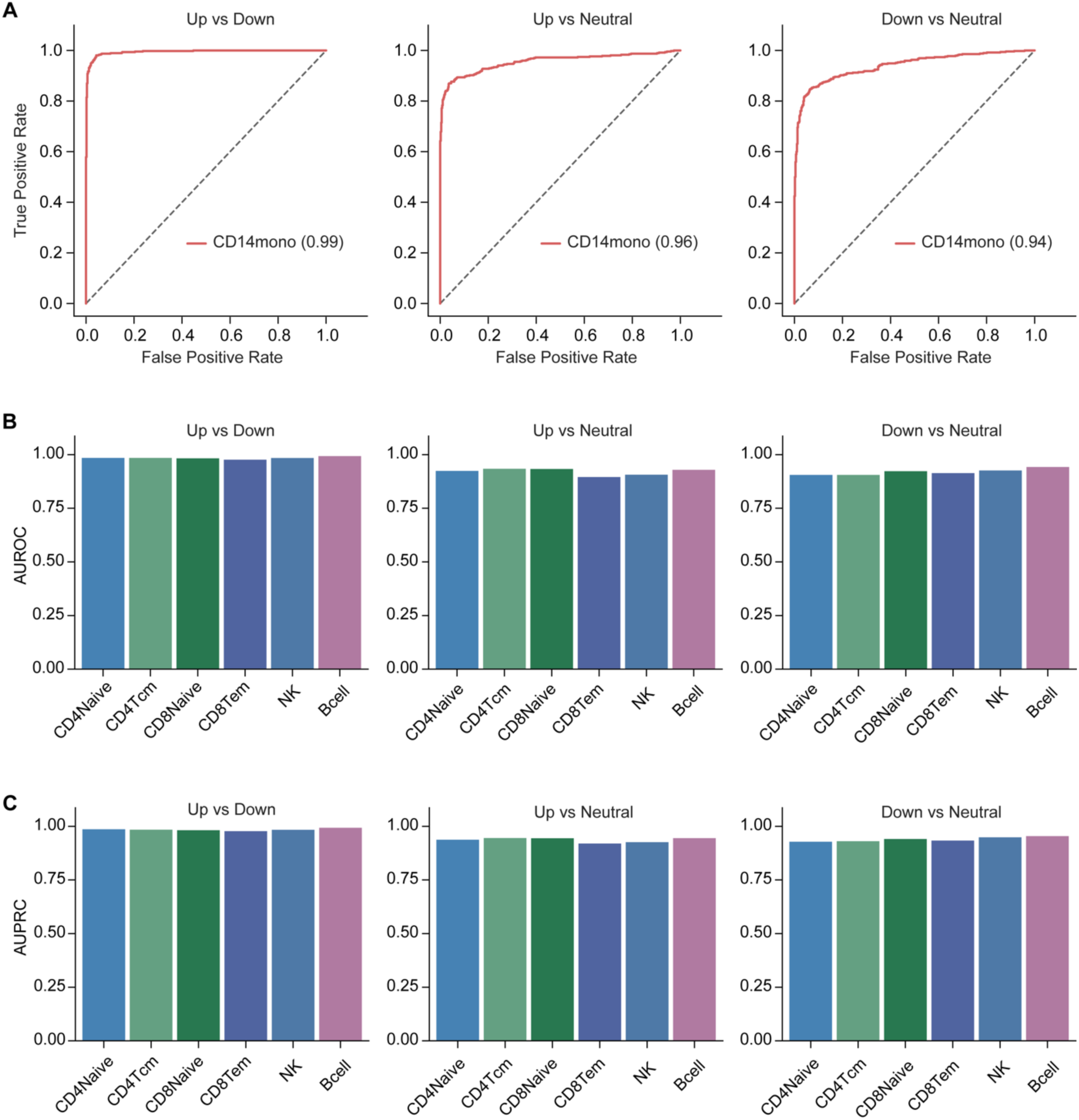
Benchmarking cross-cell-type generalization of the REG-FOCUS model. **(A)** Predictive performance (AUROC) of the REG-FOCUS model on a held-out validation set of experimentally defined effect (Up/Down) and non-effect (Neutral) variants in CD14 monocytes. **(B, C)** REG-FOCUS was applied to experimentally defined effect and non-effect variants from six additional immune cell types, and predictive performance was assessed using AUROC (B) and AUPRC (C).

**Fig. S6.**
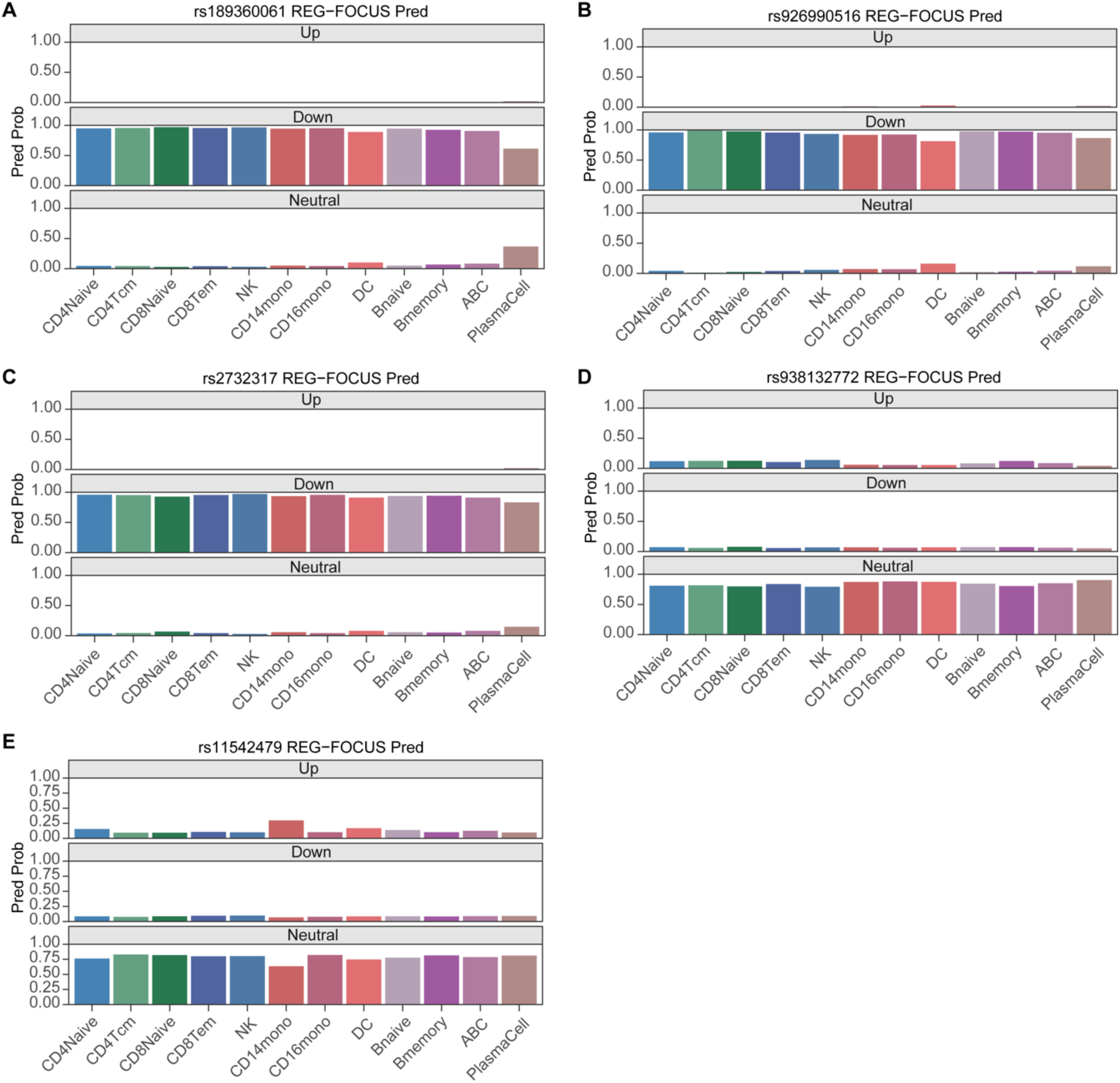
REG-FOCUS predictions for luciferase-validated regulatory variants across immune cell types. **(A-E)** REG-FOCUS predictions of the 5 variants which were validated by luciferase reporter assays across 12 immune cell types.

**Fig. S7.**
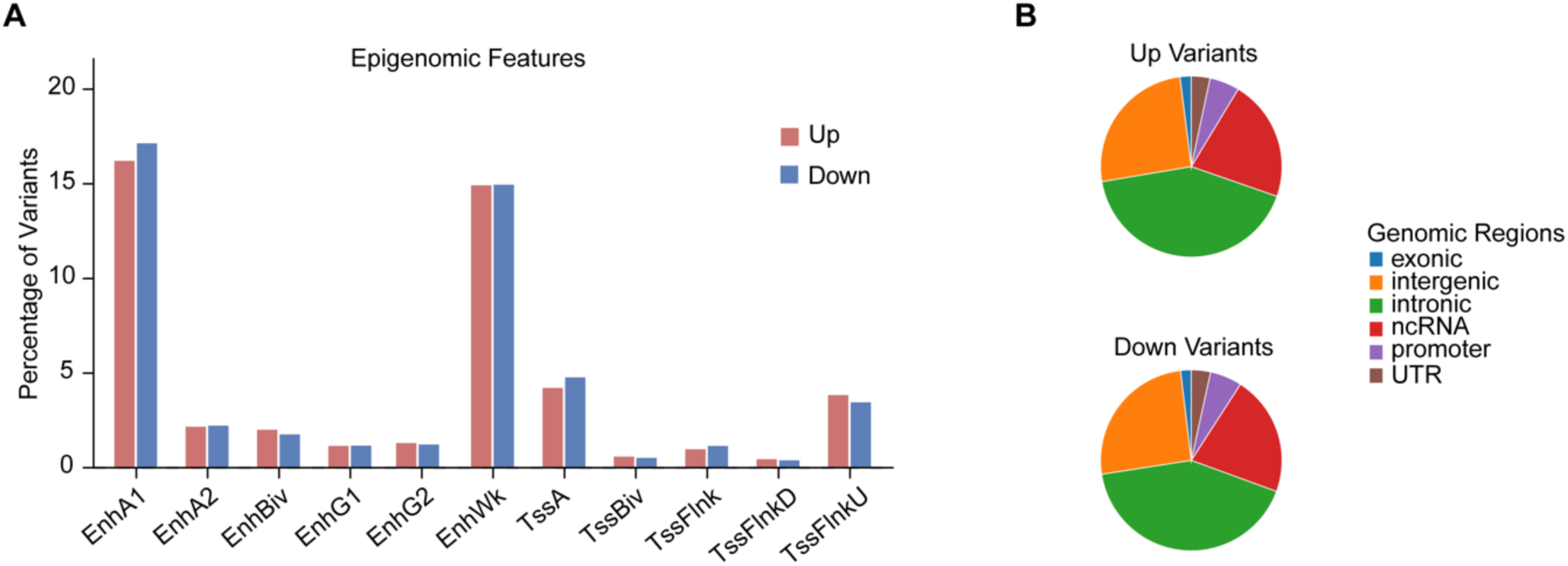
Genomic distribution of predicted effect variants. **(A)** Bar plots show the proportions of predicted effect variants across ChromHMM-defined chromatin states. **(B)** Pie charts show the proportions of effect variants across genomic regions.

**Fig. S8.**
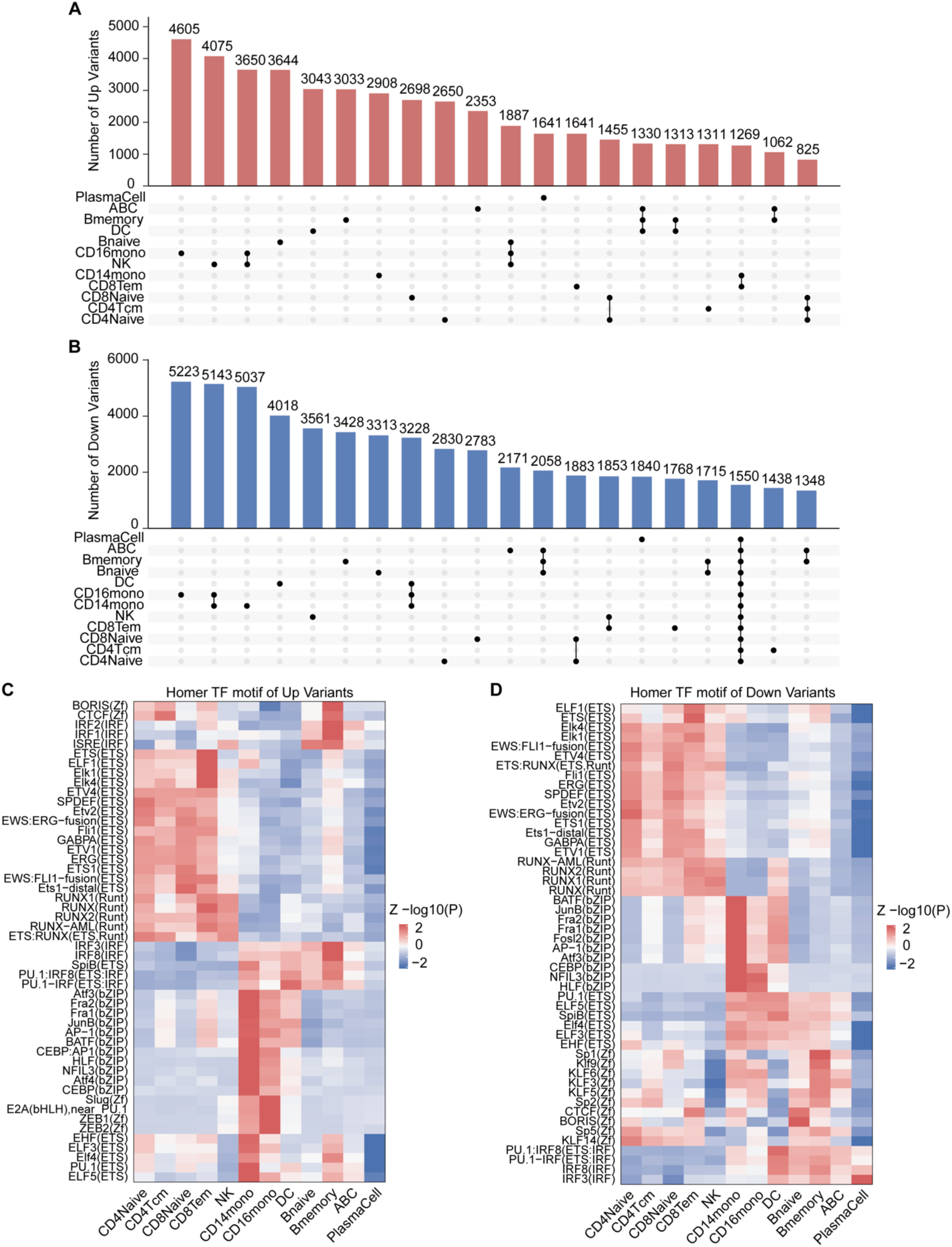
REG-FOCUS reveals cell-type-specific regulatory architectures of common variants across immune cell types. **(A, B)** UpSet plots summarize the overlap of up (A) and down (B) variants across immune cell types. **(C, D)** Heatmap of TF motif enrichment of up (C) and down (D) variants across cell types. Enrichment is represented as normalized -log10(P).

**Fig. S9.**
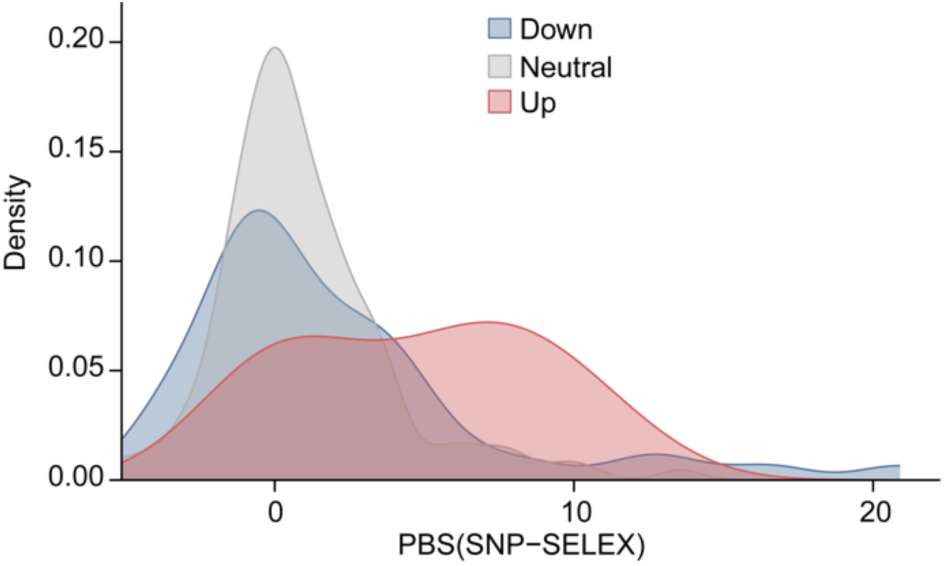
SNP-SELEX-validated TF binding effects of variants. SNP-SELEX data (experimental measurements of SNP-induced disruption of transcription factor binding) were used to evaluate the effects of variants, classified as up, down, or neutral on TF-DNA binding affinity in CD4 naive T cells.

**Fig. S10.**
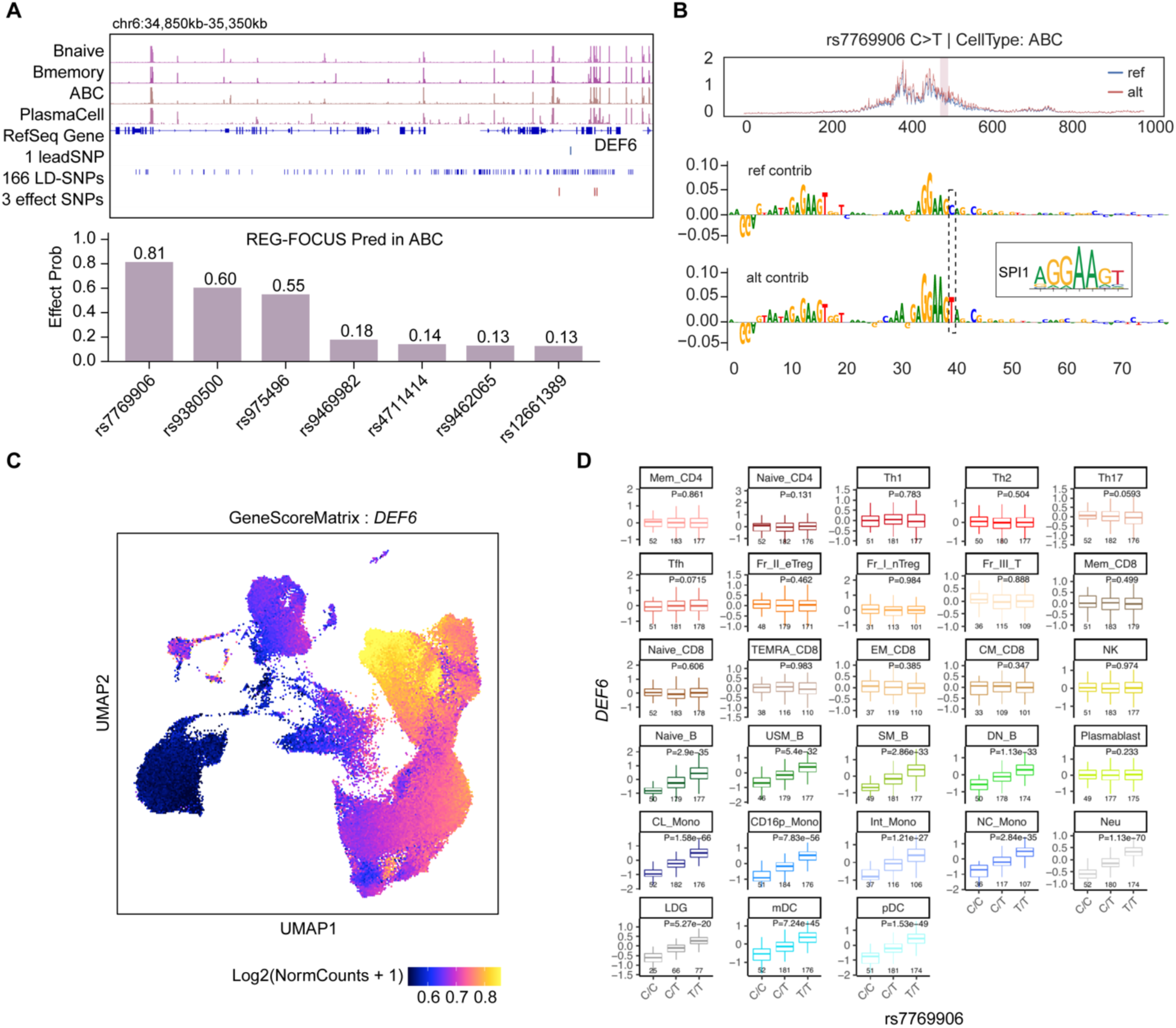
Candidate causal effect variant at the DEF6 locus in SLE GWAS. **(A)** Representation of variants screening at DEF6 locus. Tracks for chromatin accessibility, lead SNPs, SNPs in LD blocks with lead SNPs, and prioritized effect SNPs (top). REG-FOCUS predicted regulatory effects of candidate risk variants at the DEF6 locus in ABCs (bottom). **(B)** Predicted chromatin accessibility (top) and sequence contribution scores (bottom) at rs7769906 show effects on the SPI1. **(C)** UMAP embedding of *DEF6* gene activity scores across immune cell types. **(D)** eQTL associations for rs7769906 with *DEF6* across immune cell types from the ImmuNexUT dataset.

**Fig. S11.**
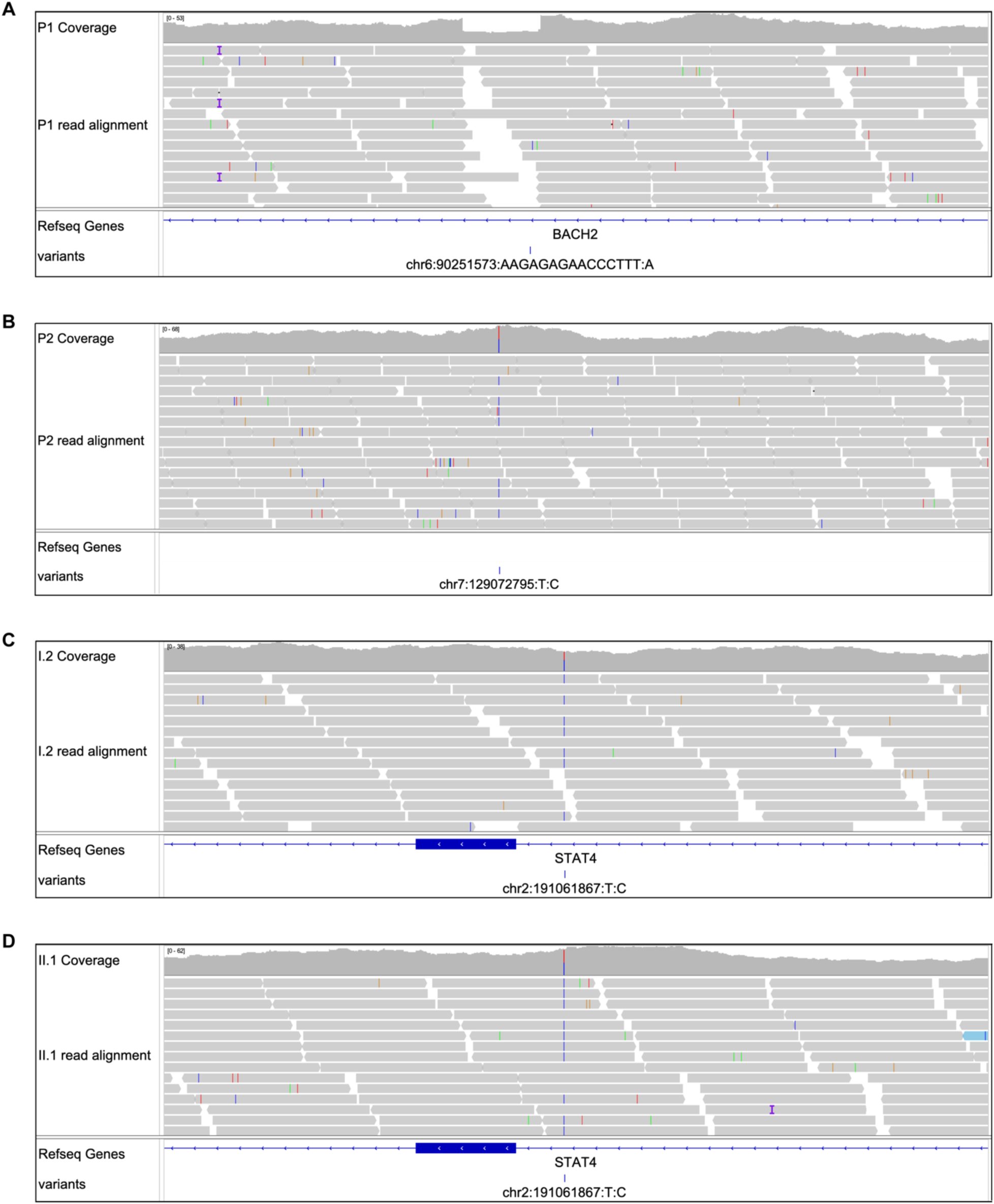
IGV browser view shows WGS identifying rare non-coding heterozygous variants in SLE patients.

**Fig. S12.**
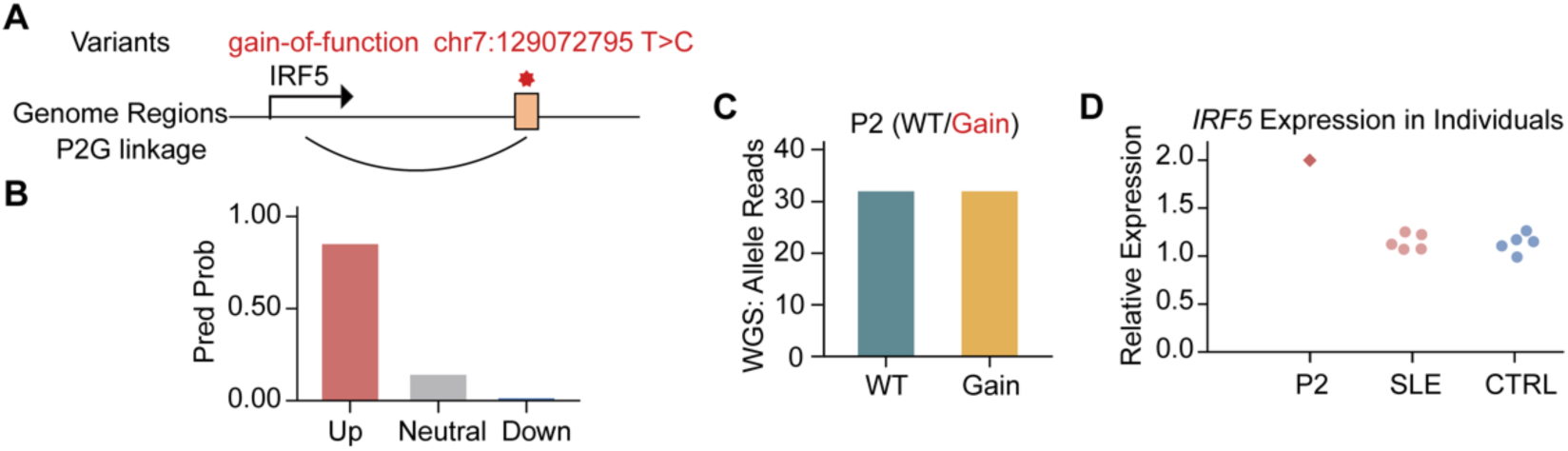
A rare gain-of-function non-coding variant at the *IRF5* locus. **(A)** Genomic location of the variant. **(B)** REG-FOCUS prediction of the variant in naive B cells. **(C)** WGS-confirmed carrier P2 of the variant. **(D)** Relative expression of *IRF5* in P2, SLE non-carriers, and healthy controls.

